# Telehealth-supported exercise/physical activity programs for knee osteoarthritis: A systematic review and meta-analysis

**DOI:** 10.1101/2023.01.18.23284720

**Authors:** Xiao-Na Xiang, Ze-Zhang Wang, Jiang-yin Zhang, Ke Li, Qi-Xu Chen, Fa-Shu Xu, Yue-Wen Zhang, Hong-Chen He, Cheng-Qi He, Si-Yi Zhu

## Abstract

Telehealth-supported program has been increasingly integrated into chronic disease management, but it is unknown if this benefit applies to populations with knee osteoarthritis. This study aimed to assess the effectiveness of telehealth-supported exercise/physical activity programs in individuals with knee osteoarthritis. A comprehensive literature search was conducted in September 2022 on randomized controlled trials investigating the effects compared to a control condition. Twenty-three studies met eligibility criteria, of which 20 studies included in meta-analysis. Telehealth-supported exercise/physical activity programs resulted in reduced pain (g = −0.39, CI −0.67 to −0.11, I^2^ = 83%), better physical activity (g = 0.13, CI 0.03 to 0.23, I^2^ = 0%) and better physical function (g = −0.51, CI −0.98 to −0.05, I^2^ = 87%). In addition, better improvements in quality of life, self-efficacy for pain, and global improvement were observed, but with non-significant improvements for self-efficacy for physical function. These findings suggested that telehealth-supported exercise/physical activity program might be an effective treatment for knee osteoarthritis. Future research with standardized components and wearables should evaluate the effects.

## 1. INTRODUCTION

Osteoarthritis is a degenerative joint disease affecting over 500 million individuals globally^1^, of whom more than 260 million have knee osteoarthritis (KOA) with notable health and socioeconomic costs^2^. The prevalence of KOA for the 60-year-old participants was 26.8%^3^, and a model forecasted that KOA will prevail in 2.37 billion individuals over 65 years and 866 million individuals over 80 years in 2100 globally^4^. The disease resulted in 9.6 million years of disability survivals (YLDs) worldwide, accounting for a 9.6% increase in the global percentage change of age-standardised YLDs between 1990-2017^5,6^. Risk factors of KOA (ie, aging, overweight, obesity) are capable of aggravating pain and disability, accelerating osteoarthritis progression, and lastly increasing the likelihood of costly joint replacement surgery^7,8^. It is estimated that the annual count of knee arthroplasty procedures will increase to 4 million by 2040 in the USA alone^9^.

As no disease-modifying drugs are available for KOA, non-pharmacologic interventions become the core treatment.^10^ In the early stage, pain and stiffness dominate symptoms over others, therefore, the management strategy focuses on the reduction of pain and the improvement of functional capacities.^11^ Clinical guidelines recommend strategies about maintaining active physical activity and exercise as first-line management for knee osteoarthritis^2,12,13^. However, gaps exist in the clinical use of active lifestyle strategies and exercise for KOA, with overusing medication and surgery^14^. Although the demand for effective intervention to cope with the need for decreased function related to an inactive lifestyle and aging seems urgent, in-person services provided by health professionals, especially physiotherapists, can be expensive regarding costs and time spent consulting an expert and the commute^15^. Besides the low time-efficiency and high cost, the lack of motivation is also a barrier since adherence to home-based exercise decreased by 94.7% after the 3 months from being discharged^16^.

Telehealth is defined as ‘the delivery and facilitation of health and health-related services including medical care, provider and patient education, health information services, and self-care via telecommunications and digital communication technologies’^17,18^. An accumulating body of evidence suggests that telehealth-supported exercise intervention has been proven as a preferable form of intervention, especially due to the ‘social distancing’ requirement imposed by the COVID-19 pandemic. Hence, the need in seeking advice or interventions via telehealth has soared^19,20^. Nonetheless, as individuals with KOA tend to be older, digital communications may have disadvantages over traditional rehabilitation due to its unclear instructions, unfamiliarity, barriers related to devices such as using a smartphone or registering on an application, and availability of Wi-Fi or cellular data.

Several reviews have attempted to confirm the efficacy of telehealth-supported exercise programs in individuals with KOA, but merely focused on the efficacy of telehealth-supported physical activity programs. Our previous meta-analysis^21^ (k = 4) indicated pain relief in patients with KOA following Internet-based rehabilitation, but the effect on physical function (g = −0.08, 95% CI: −0.27 to 0.12) was unclear due to the limited original studies included. As the intervention in the review was not inclusive (ie, programs delivered via wearables, telephone, and email were not included), the conclusion may not be robust to explain the effect of telehealth-supported exercise programs. Another meta-analysis^22^ (k = 12) using interventions via computer and virtual reality reported no improvement in physical function either (g =0.22, 95% CI: 0 to 0.43). The latest meta-analysis of 9 RCTs by Yang and colleagues^23^ indicated the telehealth-based exercise intervention was better than usual care in improving pain and able to achieve similar effects of physical function compared to in-person exercise programs (g = −0.17, 95%-CI: −0.42 to 0.08). The results of physical function are negative mainly due to the different interventions and comparators in these reviews, especially components of telehealth-supported exercise programs vary among studies. In addition, the effects on physical activity and self-efficacy in coping with symptoms and global improvement experienced by patients are not investigated.

As numerous studies have researched the effect of telehealth-based exercise/physical activity programs in KOA^24-33^, we did a systematic review and meta-analysis to investigate the effect of telehealth-supported exercise/physical activity programs on pain, physical activity, physical function, self-efficacy, quality of life and global improvement. Furthermore, the minimal important differences (MIDs) are important considerations in making clinical conclusions and are reported in this study.

## 2. RESULTS

### Study selection

A total of 14081 and an additional 13 articles were retrieved from databases. After removing duplicates, 4021 records were screened for the titles and abstracts. A total of 86 full-text articles were assessed for eligibility. Of these, 23 articles^24-48^ were included in the systematic review (see Fig. 1). List of studies excluded at the full-text screening stage with reasons is in Supplementary Table 1. Three articles^35,47,48^ were excluded for the meta-analysis due to nonstandard data reporting, resulting in 20 articles being included in the meta-analysis.

**Figure 1:**
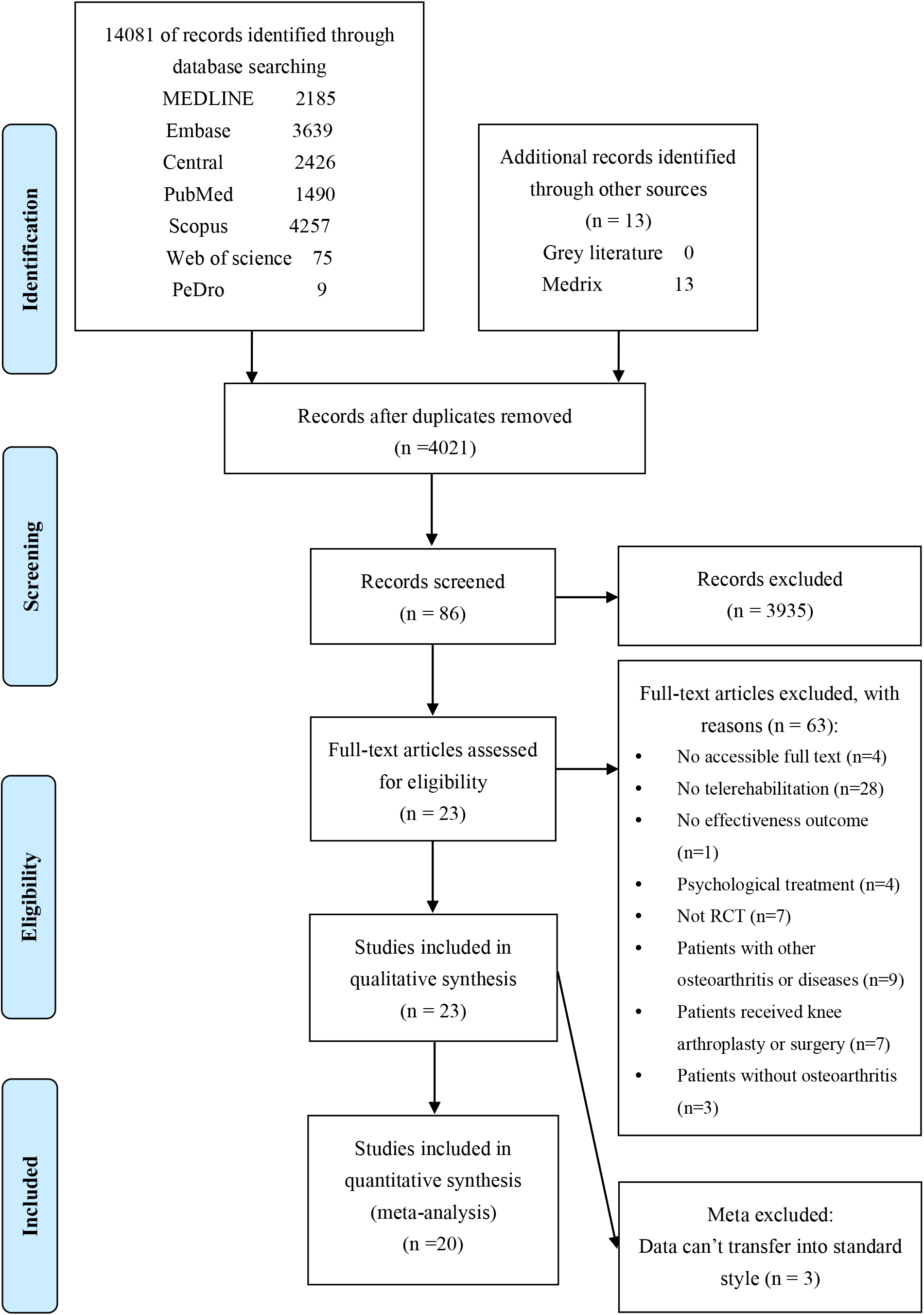
Study selection.

### Characteristics of included studies

The characteristics of all included studies are described in Table 1. A total of 23 studies involved 3824 patients, of which 60% of the participants were female, were included in the systematic review. One study^27^ observed the effect in the female population only, and others recruited participants of both sexes. Two (9%) studies were conducted in Europe, five (22%) in North America, five (22%) in Asia, nine (39%) in Australia, and two (9%) in Africa. The age of included participants was 61 (SD 3.9) years. The study period varied between 4 and 96 weeks, with seven trials (30%) performing the telehealth-based intervention for less than 3 months, six (26%) between three to six months, and 9 (39%) more than 6 months. 13 (57%) studies received telehealth-supported exercise programs, four (17%) received a physical activity program, and six (26%) received both treatments in combination. These studies tested different digital technologies, including mobile applications (k = 4), telephone (k = 4), Internet (k = 4), SMS (k = 2), and combinations (k = 9). Moreover, studies tested different additional components, such as reminder alone (k = 4), remote coaching alone (k = 3), remote monitoring alone (k = 3), combined remote reminder and monitoring (k = 2), combined remote coaching and monitoring (k = 7), with fewer studies focused on combined remote reminder and coaching (k = 1) and combined all (k = 1). For models of telehealth delivery, 8 (35%) studies used virtual contact, 5 (22%) studies used no interacting contact, 5 (22%) studies used mixed models, and one study (4%) used in-person delivery.

### Risk of bias

Risk of bias appraised with the Cochrane Collaboration’s risk of bias tool and additional PEDro scale are presented in Supplementary Fig. 1 and Supplementary Table 2. Risk of bias assessment revealed the following sources of bias: the absence of adequate random sequence generation in five (22%) studies^31,35,45,46,48^; no details of allocation concealment in five (22%) studies^29,32,35,46,47^; participants and personnel were unblinded in all studies (100%)^24-48^ limited by the intervention; the absence of the blinding of outcome assessment in nine studies (39%)^25,27,31,35,38,41,46-48^; insufficient strategies for dealing with incomplete outcome data in six (26%) studies^24,27,28,32,46,47^; no reporting bias in all studies and the presence of other bias in four studies (17%) (including insufficient power^27^ and without clinical registration^46-48^).

### Main analyses

Of these studies, 20 (87%) studies contributed data that were considered appropriate for meta-analysis. The GRADE summary of findings can be found in Supplementary Table 3. Meta-analysis results of the effects of telehealth-based exercise/physical activity programs on primary outcomes are presented in Table 2.

### Effectiveness on pain

For pain relief, there was a significant difference and a small effect size across 19 studies (n = 2512; g = −0.39; 95% CI −0.67 to −0.11; *p* = 0.0004; forest plot Fig. 2A) with high heterogeneity (*I*^*2*^=83%) for favouring the telehealth-based intervention. The calculated MID of pain was 1.3, which was lower than the reported MID (2.0 units for the Western Ontario and McMaster Universities Osteoarthritis Index (WOMAC) pain subscale).^49^ Overall, there was low-certainty evidence that the telehealth-based exercise/physical activity programs led to a small, significant, but not clinically meaningful reduction in pain.

**Figure 2:**
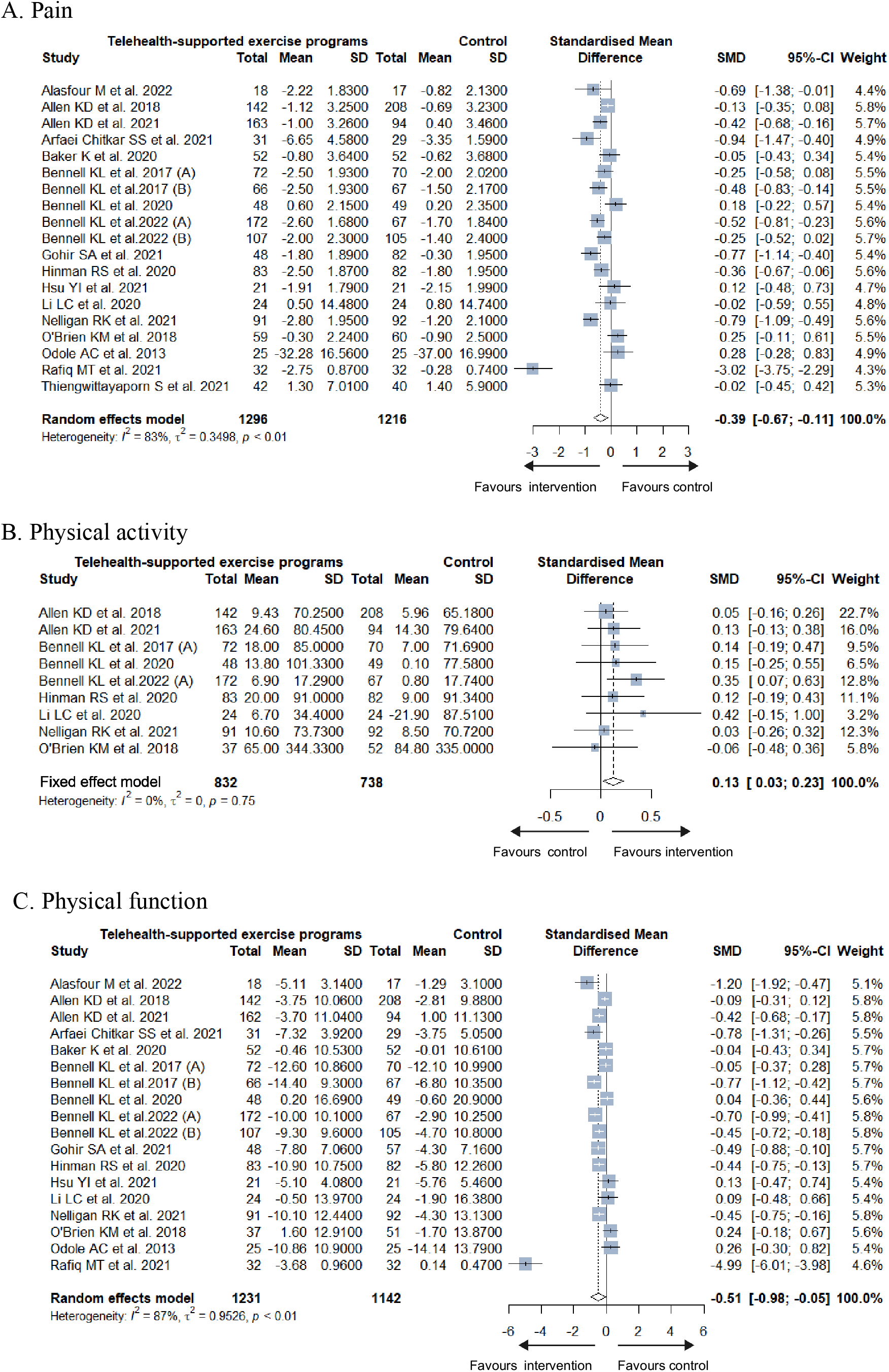
Forest plot of pain, physical activity, and physical function.

### Effectiveness on physical activity

The meta-analysis result across nine studies for physical activity favoured the telehealth-based intervention (n = 1570; g = 0.13; 95% CI 0.03 to 0.23; *p* = 0.01; forest plot Fig. 2B) with low heterogeneity (*I*^*2*^ = 0%). The calculated MID of physical activity was 9.0, which was lower than the reported MID (46.0 units for Physical Activity Scale for the Elderly).^50^ Overall, there was low-certainty evidence that the telehealth-based exercise/physical activity programs for the treatment of KOA might provide a significant, but insufficient, and not clinically meaningful improvement in physical activity.

### Effectiveness on physical function

For the improvement of physical function, the meta-analysis of 18 RCTs favoured the telehealth-based intervention with g = −0.51 (n = 2373; 95% CI −0.98 to −0.05; *p* = 0.0004; *I*^*2*^ = 87%; forest plot Fig. 2C). The calculated MID of physical function was 5.3, which was lower than the reported MID (10.1 units for the WOMAC physical function subscale).^51^ Overall, there was low-certainty evidence that telehealth-based exercise/physical activity programs might lead to a moderate, significant, but not clinically meaningful improvement in physical function.

### Effectiveness on secondary outcomes

Better secondary outcomes, such as quality of life (n = 1301; g = 0.25; 95% CI 0.14 to 0.36; *p* <0.00001; *I*^*2*^ = 5%; Supplementary Fig. 2), self-efficacy for pain (n = 1337; g = 0.72; 95% CI 0.53 to 0.91; *p* < 0.00001; *I* ^*2*^= 4%; Supplementary Fig. 2), and global improvement (n=1042; odds ratios [OR] = 2.69; 95% CI 1.41 to 5.15; *p*=0.0005; *I*^*2*^ = 79%; Supplementary Fig. 2) were observed with the intervention groups compared with control groups, but with a non-significant trend and moderate heterogeneity between studies for self-efficacy for physical function (n = 578; g = 0.14; 95% CI −0.26 to 0.53; *p* = 0.50; *I*^*2*^ = 52%; Supplementary Fig. 2). Overall, there was moderate-certainty evidence that telehealth-based exercise/physical activity programs improved the quality of life and self-efficacy for pain, and low-certainty evidence that telehealth-based intervention might provide benefit in global improvement, but not significant in self-efficacy for physical function.

### Sensitivity analyses

After removing one study^29^, the overall significant effect size for pain relief was still small with g = −0.28 (n = 2448; 95% CI −0.44 to −0.11; *p*=0.0003; Supplementary Fig. 3) and a lower heterogeneity (*I*^*2*^ = 69%), which was a robust result. Moreover, after omitting the study^29^, the overall significant effect size for physical function improvement was small with g = −0.30 (n = 2309; 95% CI −0.47 to −0.13; *p* = 0.0002; Supplementary Fig. 3) with a lower heterogeneity (*I*^*2*^ = 69%).

### Subgroup analysis

The result of the meta-regression test is listed in Supplementary Table 4. According to the World Health Organization (WHO) classification, studies were divided into (1) interventions for clients subgroup, (2) interventions for healthcare providers subgroup, and (3) interventions for clients and healthcare providers subgroup. In subgroup that included interventions for clients and healthcare providers, the effects of the telehealth-based intervention on pain (g = −0.29; 95% CI −0.49 to −0.09) and physical function (g = −0.36; 95% CI −0.63 to −0.08) were significant. However, no significant differences were found in both pain (g = −0.73; 95% CI −1.47 to 0.01) and physical function (g = −0.98; 95% CI −2.21 to 0.26) in interventions for clients subgroup. The interventions for healthcare providers subgroup showed that participants in the intervention group had no improvement in pain relief (g = 0.21; 95% CI −0.20 to 0.62) and physical function (g = 0.20; 95% CI −0.21 to 0.61) compared with the control group, either. The subgroup differences were significant in pain (*p* = 0.039) and physical function (*p* = 0.04), although heterogeneity was still significant in some of these sub-groups (*I*^*2*^>50%, Fig. 3).

**Figure 3:**
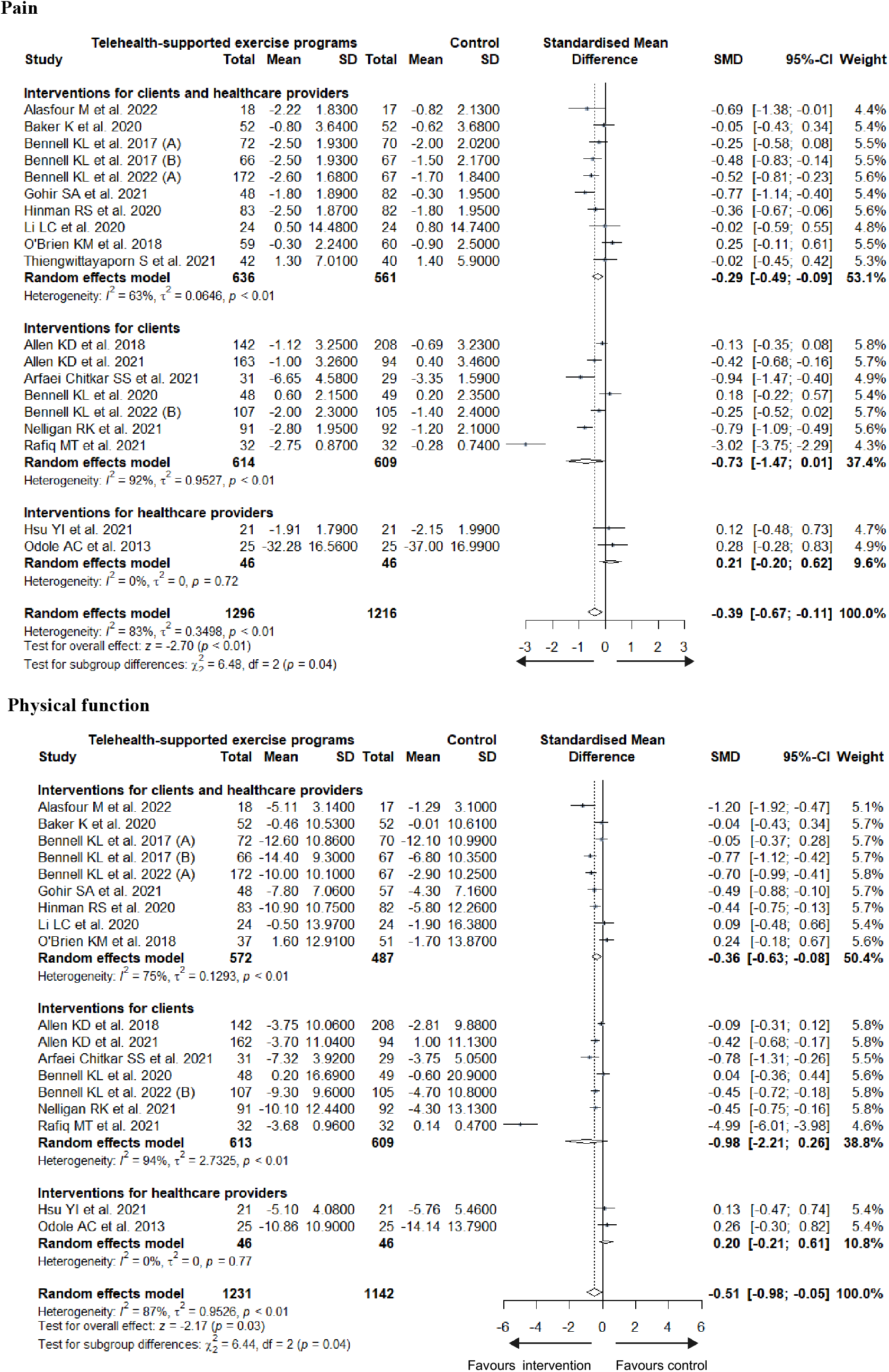
Subgroup analysis of pain and physical function based on the WHO classification.

As for the type of digital technologies in intervention group, the subgroup differences were significant in pain (*p* = 0.003) and physical function (*p* = 0.012) with high heterogeneity in mixed and SMS subgroup (*I*^*2*^ > 50%, Fig. 4). The subgroup analysis showed significant relief in pain (mobile application subgroup: g = −0.59 (95% CI −1.01 to −0.16); Internet subgroup: g = −0.25 (95% CI −0.43 to −0.07); and mixed type: g = −0.29 (95% CI −0.57 to −0.01)), and significant improvement in physical function (mobile application subgroup: g = −0.73 (95% CI −1.10 to −0.36); Internet subgroup: g = −0.42 (95% CI −0.80 to −0.04); and mixed type: g = −0.28 (95% CI −0.54 to −0.02)). Nonetheless, interventions with telephone and SMS subgroup showed both non-significant results in pain (g = −0.08; 95% CI −0.34 to 0.18) and g = −1.41; 95% CI −4.54 to 1.73), respectively) and physical function (g = 0.01; 95% CI −0.22 to 0.23) and g = −2.45; 95% CI −7.39 to 2.48, respectively).

**Figure 4:**
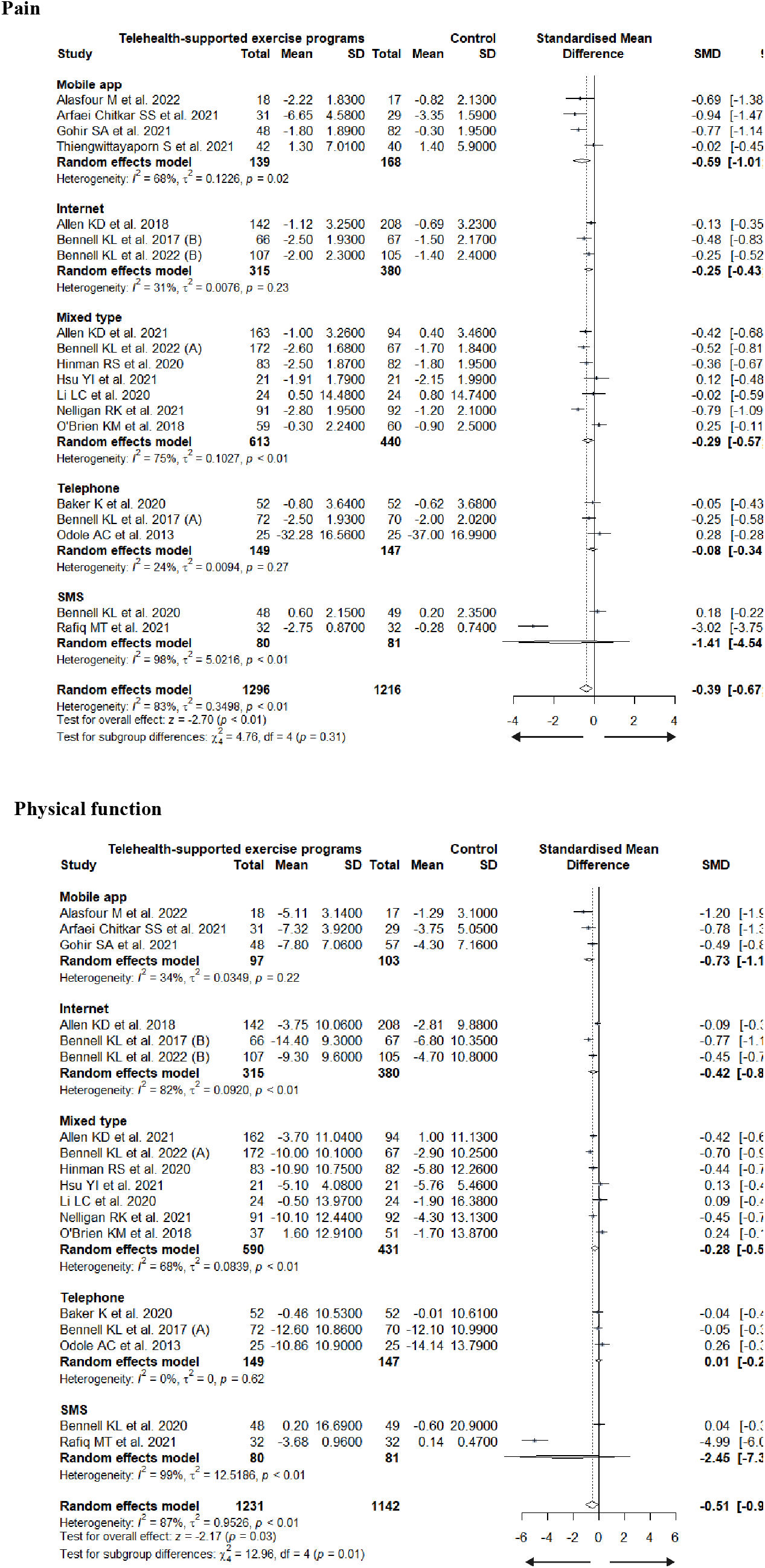
Subgroup analysis of pain and physical function based on the type of digital technology.

The control treatments in included studies were divided into active controls (ie, exercise, physical therapy, and self-management) or inactive controls (ie, education, usual care, and waitlist) with subgroup differences (pain *p* = 0.021; physical function *p* = 0.065, Fig. 5). Compared with the active control groups, no statistically significant difference in pain (g = −0.08; 95% CI −0.21 to 0.05) and physical function (g=−0.07; 95% CI −0.21 to 0.07) was determined. Compared with inactive control groups, statistically significant pain reduction (g = −0.63; 95% CI −1.08 to −0.18) and function improvement (g = −0.79; 95% CI −1.54 to −0.03) were found in the intervention group.

**Figure 5:**
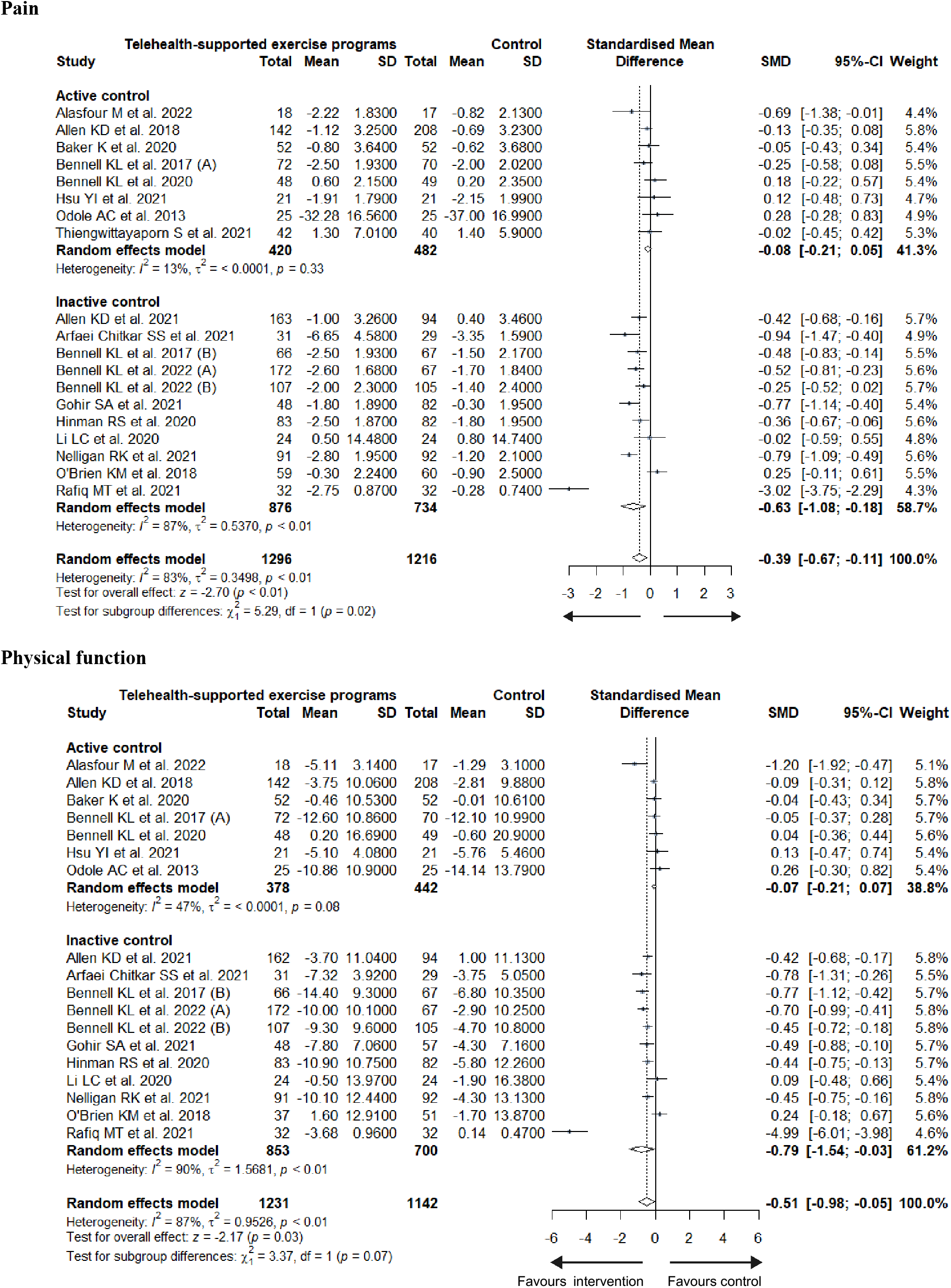
Subgroup analysis of pain and physical function based on the control group.

### Publication bias

Visual assessment of funnel plots for studies reporting pain relief, and improvement in physical activity and function did not suggest publication bias (see Supplementary Fig. 4) and these were supported by Egger’s regression test for plot (see Table 2).

## 3. DISCUSSION

We systematically reviewed the effectiveness of telehealth-supported exercise/physical activity programs on pain, physical activity, physical function, self-efficacy, quality of life, and global improvement among individuals with KOA. Overall, this review found low-certainty evidence that telehealth-supported programs might show a small benefit on pain, a very small effect on physical activity, and a moderated effect on physical function, although these changes of primary outcomes were not significant in clinical difference. Additionally, this review found moderate-certainty evidence that telehealth-supported exercise/physical activity programs might provide a statistical improvement in quality of life and self-efficacy for pain, and low-certainty evidence that telehealth-supported interventions might increase global improvement. But we found low-certainty evidence suggesting that telehealth-supported exercise/physical activity programs made no significant improvement in self-efficacy for physical function. Of studies that were categorized as interventions for clients and healthcare providers according to WHO classification (v1.0)^52^, the benefits in pain relief and physical function improvement were found. We also found that digital technology affected the results, and positive effects were only delivered by mobile applications, Internet, and mixed type. Additionally, our subgroup analysis of control interventions indicated that telehealth-supported exercise/physical activity programs had a greater increase in pain and function than inactive intervention did. Although we thought the reminder, monitoring, and coaching should be important components of telehealth-supported programs, no significant subgroup difference was found. These findings suggest a definite role for telehealth-supported exercise/physical activity programs in the management of KOA, and formative research is required to better understand the trend in telehealth technology, and the component of telehealth-supported programs.

Although we reported the benefits of telehealth-supported exercise/physical activity programs, no improvements were determined in the self-efficacy for physical function. Self-efficacy for physical function represented the perceived capability to improve physical function, which is subjective and causally influences outcome expectancies, but not vice versa^53^. In our meta-analysis, more objective result by scales indicated that improvement in physical function might exist. Nonetheless, the inconsistency with the results of self-efficacy for physical function indicated participants who received the telehealth-supported exercise/physical activity programs might have few motivations in increasing function. This condition might reflect the influence of previous studies. Only two meta-analyses^22,23^ reported the effects of telehealth-based exercise programs on physical function, and reported negative conclusions. These dispiriting results might decrease the motivation for physical function. Moreover, exercise is one type of planned, structured, and repetitive physical activity^54^, it is more essential to determine the effect of telehealth-based physical activity programs. Most telehealth-based programs contained strategies about improving activities of sports, daily living, occupation, leisure, active transportation, and energy expenditure above resting levels, that aimed at improving physical activity. Therefore, our meta-analysis included studies involving telehealth-based exercise or physical activity interventions and changed the conclusion about physical function.

Multiple factors affected the performance of telehealth-based interventions and resulted in high heterogeneity. We did sensitive and subgroup analysis to explain the source of heterogeneity, and we found the study of Rafiq and the colleagues increased the heterogeneity because their results of telehealth-based programs reflected huge improvements. The improvements might be caused by telehealth-based strengthening exercise programs and the reminders to clients that required clients to complete the exercise session^29^. Reminders may increase the effect of telehealth programs by improving adherence to telemedicine potentially^55^. Nonetheless, our review did not identify evidence of heterogeneity and effect when comparing whether the telehealth-based programs with a reminder, coaching, and monitoring or not. Consultations or coaching, as a professional continuous service, might play an important role in transmitting health information and providing decision support^56^. Palamara and colleagues^57^ reported an improvement in well-being among female surgery residents who received vital coaching over a web-based video platform. Nonetheless, Sieczkowska et al.^58^ revealed the available evidence is not of sufficient quality to support the use of self-reported health coaching as a health care intervention for weight loss. Although face-to-face clinician contact might be considered crucial for the delivery of available specialist knowledge, consultations or coaching between remote client and healthcare provider^59^, targeted alerts and reminders to clients^60^, and remote monitoring of client^61^ could still be controversial. More studies with strict design should focus on the effect of remote consultations, coaching, reminders, and monitoring to standardise the format components of telehealth-based exercise/physical activity programs and understand the clinical importance of telehealth-supported exercise/physical activity programs.

Besides the components in programs, we found the targeted primary users, digital technology, and comparators were more important in our study. We have classified the telehealth-based programs with the WHO classification (v1.0)^52^, which is a good and useful tool for identifying the characteristics of telemedicine. Our results emphasized the interactions between clients and health providers contributed to improvement of pain and physical function. This WHO classification of digital health interventions matched the trend of digital technologies, and might will be developed to constrain smart technologies, such as robotics-assisted training, physical therapy device, and exercise sensors via the Internet of things^62^. As for technology delivered the telehealth-based programs, only two studies^25,43^ used wearables for personal data tracking, which was consistent with the conclusion that nonoperative osteoarthritis patients were generally unreceptive to using wearable technologies^63^. Most recent trials employed mobile applications and the Internet, that reflect personal telemedicine devices might be the trend of delivery methods, recent societal and technological advances. It is easy to understand that the control interventions may be a potential source of heterogeneity. We found patients in telehealth-based exercise/physical activity programs gained equivalent effects on pain reduction and physical function with active controls, such as physical therapy and face-to-face exercise, and superior effects than inactive controls, such as usual care and education. Therefore, patients might be inspired by the substantial benefits of telehealth achieved by results with convenient access and at low cost under the condition of constrained medical resources^64^.

Our results updated evidence based on previous systematic review and meta-analysis by Yang et al.^23^, Chen et al.^22^, and McHugh and co-authors^65^. Yang et al.^23^ and Chen et al.^22^ included nine studies about telehealth-based exercise intervention and 12 RCTs about technology-supported exercise programs reporting up to June, 2021 and August, 2020, respectively. We focused more on the telehealth-based physical activity intervention, and determined the small, but positive effect of telehealth-based exercise/physical activity programs on improvement in physical activity. Our results of physical function and quality of life were positive and in contrast to these two meta-analyses, which might be due to more studies appeared recently. Our subgroup about digital technology was similar to the review by Yang and co-authors^23^ that also analysed the delivery technology of telehealth-based programs, reporting seven of nine studies used websites or telephones. Although the review by McHugh and co-authors^65^ analyzed the effectiveness of remote exercise programs in reducing knee pain about varied comparators, they only described the difference between inactive comparators and active comparators without quantitative analysis, while we completed an accurate analysis. Moreover, we provided the MIDs of pain, physical activity, and physical function, although the values were below the values of clinical importance.

Although the effectiveness of telehealth-supported exercise/physical activity programs for KOA was proved beneficial in this study, the variation in remote technologies and management of data are still challenges for popularizing and applying for the telehealth-supported exercise/physical activity program as a first-line therapy. Smart wearables, such as smartwatches and smart suits, seem to be the current innovation drivers, that are gradually widely used devices worldwide. Although patients used to be unreceptive to using wearable technologies, the development of this technology increases rapidly, especially for direct-to-consumer wearable tracking technologies^66^, and might increase the acceptance of wearable technologies, and help to facilitate the consistency in remote technologies. The adaption of remote technologies and health information, the fluency of information transmission, the quality and consistency of responses to clients based on the interpretation of individual data, and the cost of exploring new technologies^67^ gain more and more focus, and could boost the bloom of telehealth-supported programs.

The key strengths of this study included the investigation of telehealth-supported exercise/physical activity programs on physical activity, and physical function with self-efficacy, to better illustrate the effects of telehealth-supported exercise/physical activity programs. Another strength was the comprehensive systematic component of the literature review, which led to new evidence and identified significant factors that, if concentrated in future research, could facilitate a better understanding and development of telehealth-supported exercise/physical activity programs. There are some limitations in our study. Firstly, although the source of heterogeneity was partially identified, there still was some heterogeneity in the included RCTs for insidious reasons. Secondly, the long-term benefits of telehealth-supported exercise/physical activity program interventions were unknown, as few studies completed a long-term assessment after interventions for more than 3 months. Lastly, it is difficult for researchers to blind allocation for participants and personnel, and this might become another factor contributing to the high heterogeneity.

## 4. CONCLUSION

This systematic review found low certainty evidence that telehealth-supported exercise/physical activity programs for the treatment of KOA might provide a very small to moderate, and not clinically meaningful improvement in pain intensity, physical activity, and physical function. Additionally, moderate-certainty evidence suggested improvement in quality of life and self-efficacy for pain, and low-certainty evidence suggested an increase in global improvement. But low-certainty evidence suggested that telehealth-supported exercise/physical activity programs made no significant improvement in self-efficacy for physical function. Future research needs to standardize the components of telehealth-supported exercise/physical activity programs and focus more on wearable technologies to provide a high-level of evidence to support clinical practice.

## 5. METHODS

The review protocol was registered with PROSPERO (CRD42022359658) and was performed according to the PRISMA recommendations (Preferred Reporting Items for Systematic Reviews and Meta-analyses)^68^.

### Search strategy

In this systematic review and meta-analysis, we searched Embase (via OVID platform), MEDLINE (via OVID platform), CENTRAL (via the Cochrane Library), Web of Science, PubMed, Scopus, and the Physiotherapy Evidence Database from inception to September 2022, for randomized controlled trials (RCTs) published in English language peer-reviewed journals. The specialist register GreyNet (http://www.greynet.org/) and medRxiv (https://www.medrxiv.org/) were searched for grey literature. The reference lists in studies included for full-text screening were also hand-searched to identify potentially relevant articles. We included studies that intervention group involved telehealth-supported exercise/physical activity programs (including the use of SMS, videos, email, Internet, and applications with or without wearable devices) as the main treatment for KOA patients. The comparator groups included patients who did not receive a telehealth-supported exercise/physical activity intervention. The key search terms included “telerehabilitation”, “exercise or physical activity”, “knee osteoarthritis”, and “randomized controlled trial”. We developed a search strategy via OvidSP and the full search strategy is listed in the Supplementary Table 5.

### Eligibility criteria

Studies were included if they met the following inclusion criteria:

#### Population

participants regardless of age with a diagnosis of KOA.

#### Intervention

telehealth-supported exercise/physical activity programs delivered by SMS, videos, email, Internet, or applications combined with wearable devices as major intervention.

#### Comparator

included telehealth-supported programs without exercise or physical activity, or waiting list or non-telemedicine interventions (ie, usual care, conventional exercise programs, patient education, etc.). Control interventions containing telehealth-supported programs with education were not excluded, because achievable exercise/physical activity strategy was omitted.

#### Outcomes

pain measured by the WOMAC pain subscale, visual analogue scale, Knee Injury and Osteoarthritis Outcome Score (KOOS) pain subscale, or Numeric Pain Rating Scale, physical activity measured by PASE, time spent in daily moderate-to-vigorous physical activity, International Physical Activity Questionnaire, or International Physical Exercise Questionnaire, and physical function measured by WOMAC function subscale or KOOS function subscale. The secondary outcomes were quality of life measured by KOOS Quality of Life subscale or Assessment of Quality of Life, self-efficacy measured by Arthritis Self-Efficacy Scale pain and function subscale, and overall global improvement.

#### Study types

randomised controlled trials.

Studies were excluded if they were cases, letters, comments, trial protocols, conference articles, editorials, reviews, or practice guidelines. Articles were excluded if they did not investigate the changes in pain, physical activity, physical function, quality of life, self-efficacy, or global improvement. In addition, studies with other arthritis diseases and unclear statistical analysis or reporting of results were also excluded.

### Selection criteria

Two authors (XNX and ZZW) independently performed the initial screening and study selection according to titles and abstracts. Any disagreements were resolved by a discussion under the guidance of a third reviewer (SYZ). Both reviewers (XNX and ZZW) read the full texts of articles. Conflicts over inclusion were adjudicated by a third reviewer (SYZ).

### Data extraction

Our primary outcomes were pain, physical activity, and physical function. Our secondary outcomes included the quality of life, self-efficacy for pain and function, and global improvement. Two independent authors (ZZW and JYZ) extracted data with a standardized data template. The following data were extracted: country, sampling methodology, sample size, participants’ age, detailed telehealth-supported exercise and comparator, time point, the WHO classification of digital health^52^, and statistical reporting of mentioned outcomes. If SDs were missing for continuous data, other statistics (ie, 95% confidence interval, standard errors, *T/F/p*-values, etc.) were used for the calculation of SD via the calculator tool from Review Manager, version 5.4 (Nordic Cochrane Centre, Cochrane Collaboration). Disagreements between the two reviewers were resolved by consensus, and if necessary, by consultation with a third reviewer (XNX).

### Data analysis

Data analysis was performed with Review Manager and R (version 4.2.1). For the continuous variable, Hedges’ g (g) with 95% confidence interval (CI) was used for the same measurement across studies for the specified outcomes or not, respectively. Heterogeneity was assessed using Cochrane *Q* statistic (significance level at *p* < 0.05) and quantified with *I*^*2*^ (significance level at *I*^*2*^ > 50%)^69,70^. Random effects were used if the *Q* or *I*^*2*^ value was statistically significant, or a small number of studies were analysed. Otherwise, fixed effects were used. Egger’s regression test and funnel plot of the primary outcomes were used to assess the possibility of publication bias^71^. If the test for asymmetry was significant, trim-and-fill method was used to adjust for possible bias by obtaining an estimation of the pooled effect when accounting for missing studies. A sensitivity analysis was performed for primary outcomes to ensure robustness^72^.

A meta-regression test was used for determining the factors which might explain the heterogeneity. The subgroup analysis of the primary outcomes was reported based on: (1) the WHO classification of digital health,^52^ (2) the category of telehealth technologies, and (3) the type of treatment (active or inactive interventions) received by control groups. The Hedges’ g cut-off points of 0.20, 0.50, and 0.80 can be considered to represent a small, moderate, and large effect, respectively. *P* value < 0.05 was considered statistically significant. MIDs of primary outcomes were calculated on the Hedges’ g and pooled represented SDs (pooled from the intervention and control groups in a trial that used the scale)^73^, and compared with reported MIDs. The anchor-based estimates were used if there was no established MID^74^.

### Quality assessment

The Cochrane Collaboration’s risk of bias tool was used to assess bias, we assessed each domain, as follows: selection bias, performance bias, detection bias, attrition bias, reporting bias, and other sources of bias. Each component was recorded as low, unclear, or high risk of bias. Additionally, the quality of included studies was measured by the Physiotherapy Evidence Database (PEDro) scale for preciseness.

### Quality of evidence assessment

GRADE (Grading of Recommendations Assessment, Development, and Evaluation) approach was used to evaluate the certainty of evidence for each outcome. For each of the outcomes, the overall certainty of evidence for each outcome was graded as high, moderate, low, or very low, and evidence was downgraded by 1 level from high quality for each serious problem found in the domains of risk of bias, inconsistency (substantial heterogeneity: *I*^*2*^ > 50%), indirectness, imprecision (such as small sample size), and publication bias^75^.

## Data Availability

All data produced in the present work are contained in the manuscript

## Author Contributions

CQH, SYZ, and XNX were responsible for concept, study design, and search strategy for this review. XNX and ZZW did all database searching and collating of results. XXN, ZZW, JYZ and SYZ did the article screening, data extraction, and critical appraisal. All authors contributed to conflict resolution during screening. SYZ and XNX were responsible for data curation. XNX, KL, QXC, YWZ and FSX contributed to data analysis and data interpretation. XNX and SYZ drafted the manuscript. All authors contributed to reviewing and editing of the final manuscript.

## Acknowledgments

We thank the Qing-Yang Shi in West China Hospital for methodology assistance.

## Data sharing

All data for this review and meta-analysis were obtained from primary literature. Data extracted and derived data will be made available on reasonable request from the first or corresponding author.

